# Reliability of novel centre of pressure measures of quiet standing balance in people with chronic stroke

**DOI:** 10.1101/2022.11.29.22282901

**Authors:** David Jagroop, Raabeae Aryan, Alison Schinkel-Ivy, Avril Mansfield

## Abstract

**Background:** People with stroke often have asymmetric motor impairment. Investigating asymmetries in, and dynamic properties of, centre of pressure movement during quiet standing can inform how well balance is controlled.

**Research question:** What are the test-retest reliabilities of novel measures of quiet standing balance control in people with chronic stroke?

**Methods:** Twenty people with chronic stroke (>6 months post-stroke), who were able to stand for at least 30 seconds without support, were recruited. Participants completed two 30-second quiet standing trials in a standardized position. Novel measures of quiet standing balance control included: symmetry of variability in centre of pressure displacement and velocity, between-limb synchronization, and sample entropy. Root mean square of centre of pressure displacement and velocity in the antero-posterior and medio-lateral directions were also calculated. Intraclass correlation coefficients (ICCs) were used to determine test-retest reliability, and Bland-Altman plots were created to examine proportional biases.

**Results:** ICC_3,2_ were between 0.79 and 0.95 for all variables, indicating ‘good’ to ‘excellent’ reliability (>0.75). However, ICC_3,1_ for symmetry indices and between-limb synchronization were <0.75. Bland-Altman plots revealed possible proportional biases for root mean square of medio-lateral centre of pressure displacement and velocity and between-limb synchronization, with larger between-trial differences for participants with worse values.

**Significance:** These findings suggest that centre of pressure measures extracted from a single 30-second quiet standing trial may have sufficient reliability for some research studies in chronic stroke. However, for clinical applications, the average of at least two trials may be required.

## INTRODUCTION

People with chronic stroke (>6 months post-stroke) typically have sensorimotor deficits, which impair balance control and increase the risk of falls [1]. Consequently, improving balance control is a major focus in stroke rehabilitation, as it enables safe discharge. Standardized balance measures are essential in clinical settings to identify specific deficits that should be targeted with rehabilitation, and to track change over time [2]. Performance-based observational rating scales, such as the Berg Balance Scale [3], are frequently used to assess balance in clinical practice [4]. However, the assessment scores do not inform compensatory strategies used to complete tasks within the scale, and in turn, do not inform underlying dyscontrol that could potentially increase the risk of falling or be targeted with rehabilitation [5].

When standing still, the centre of pressure (COP) moves to control postural sway [6]. Therefore, examining COP movement during quiet standing can inform how well balance is controlled. Previous studies have established test-retest reliability of quiet standing balance control measures in people with chronic stroke, such as area covered by the COP [7,8], mean COP velocity [7,8], and variability in COP displacement [9] and velocity [7]. These measures consider COP movement under both feet combined; however, since stroke often results in asymmetric motor impairment, it may be beneficial to assess how each limb contributes to balance control [5]. In particular, correlation between COPs under individual limbs may reveal the extent to which the limbs work together in a synchronized manner to control balance [10,11], and symmetry of COP fluctuations under both limbs may reveal how much each limb contributes to balance control [12,13]. Worse between-limb synchrony and greater asymmetry in contribution of the limbs to balance control predict increased risk of falls after discharge from stroke rehabilitation [14]. Examining dynamic properties of the COP may also provide additional insights into balance control post-stroke [15]. In particular, sample entropy of the COP may inform attentional investment in balance control [16], which may be a useful metric to inform stroke rehabilitation. Only one study has aimed to establish reliability of between-limb synchronization in people with stroke [9]; however, this study included people with sub-acute and chronic stroke, so the reliability of between-limb synchronization has not been established in people with chronic stroke, specifically. To our knowledge, no previous studies have aimed to establish the test-retest reliability of COP symmetry or sample entropy post-stroke.

The purpose of this study was to determine the test-retest reliabilities of novel measures of quiet standing balance control in people with chronic stroke, namely: symmetry of COP fluctuations, between-limb synchronization, and sample entropy. To compare to previous work and to inform our symmetry indices, we also included four other quiet standing balance measures in the analysis: root mean square (RMS) of COP displacement and velocity in the antero-posterior (AP) and medio-lateral (ML) directions.

## METHODS

### Participants

Participants were people with chronic stroke (>6 months post-stroke) who participated in a previous clinical trial that aimed to determine the effect of reactive balance training on falls in daily life [17,18]. To be eligible for the clinical trial, participants had to be able to stand for at least 30 seconds without support, and tolerate at least 10 balance perturbations using a lean-and-release system. Participants were excluded if they: had any other neurological condition; had lower-extremity amputation; were unable to understand instructions in English; had significant illness or surgery in the past 6 months; had osteoporosis with a history of fracture; had poorly controlled diabetes or hypertension; had contraindications to physical exercise; were undergoing physiotherapy for balance/mobility problems at the time of the study; or received reactive balance training in the year before enrolment. A sub-set of participants from this larger study completed additional data collection in a biomechanics laboratory. To be eligible for this additional data collection, participants had to be able to walk for at least 10 metres without a gait aid or physical support. The study was approved by our institution’s research ethics, and participants provided written informed consent to participate.

Participant age, sex, and date of stroke were obtained from participants at enrolment into the clinical trial. Participant height, body mass, National Institutes of Health Stroke Scale [19] scores, more-affected side of the body, and Chedoke-McMaster Stroke Assessment [20] leg and foot scores were measured at the pre-intervention assessment for the clinical trial. The Berg Balance Scale [3] and Timed-Up and Go [21] scores were obtained from either the pre- or post-intervention assessment for each participant, as appropriate (see Procedures section). Participant characteristics are presented in Table 1.

**Table 1:** Participant characteristics.

|  | Mean or count | Standard deviation | Minimum | Maximum |
| --- | --- | --- | --- | --- |
| Age (years) | 64.0 | 9.5 | 43.0 | 79.0 |
| Sex |  |  |  |  |
| Men | 6 |  |  |  |
| Women | 14 |  |  |  |
| Height (cm) | 169.2 | 9.0 | 148.6 | 187.0 |
| Body mass (kg) | 76.5 | 12.8 | 60.3 | 110.9 |
| Time post-stroke (years) | 4.4 | 3.7 | 0.6 | 14.4 |
| NIH-SS (score) | 3.7 | 3.3 | 0.0 | 13.0 |
| More affected side of the body |  |  |  |  |
| Left | 12 |  |  |  |
| Right | 8 |  |  |  |
| CMSA leg (score out of 7) | 5.6 | 1.2 | 3.0 | 7.0 |
| CMSA foot (score out of 7 ) | 5.1 | 1.3 | 2.0 | 7.0 |
| BBS (score out of 56) | 52.9 | 4.2 | 42.0 | 56.0 |
| Timed Up and Go (s) | 13.2 | 6.9 | 5.6 | 32.3 |

NIH-SS: National Institutes of Health Stroke Scale; CMSA: Chedoke-McMaster Stroke Assessment; BBS: Berg Balance Scale.

### Procedures

Data collection for quiet standing balance trials occurred either before (18 participants) or after (2 participants) completing the study interventions. Participants completed 2 quiet standing trials; in between each trial, participants completed a trial of over ground walking at a comfortable speed (data from walking trials are not presented in the current paper). Therefore, there was approximately 2 minutes break between each quiet standing trial. During the quiet standing trials, participants stood in a standardized position [22], with one foot on each of two side-by-side force plates (Advanced Medical Technology Inc., Watertown, Massachusetts, USA). Participants were instructed to stand as still as possible, looking straight ahead, for 30 seconds. Ground reaction forces and moments were collected at 250 Hz and stored for offline processing.

### Data processing

Data were processed using MATLAB (R2020b, The Mathworks Inc., Natick, Massachusetts, USA). Raw ground reaction forces and moments were low-pass filtered at 10Hz using a 2^nd^ order zero phase lag Butterworth filter. The antero-posterior (AP) and medio-lateral (ML) centres of pressure (COP) under each limb separately, and under both limbs combined, were calculated from the forces and moments. The root mean square (RMS) of AP and ML COP position and velocity were calculated under each limb separately and both limbs combined. Two symmetry indices were calculated as RMS of AP COP displacement and velocity under the more affected limb, divided by the sum of RMS of AP COP displacement and velocity under both limbs. Between-limb synchronization was calculated as the cross-correlation coefficient between AP COPs under each limb separately at zero lag. Symmetry indices and between-limb synchronization were not calculated using ML COPs as ML sway is primarily controlled by loading and unloading the limbs, rather than through changes in individual-limb ML COP [5,10]. The resultant COP signal was normalized by dividing by its standard deviation, and sample entropy was calculated from this normalized resultant signal using a MATLAB routine from PhysioNet [23] with the optimal input parameters determined to be M=4 and r=0.015 [16,24].

### Data analysis

Data analysis was conducted using SAS (Version 9.2, SAS Institute, Cary, North Carolina, USA). Intraclass correlation coefficients (ICCs) were calculated using the intracc macro [25]. Both ICC_3,1_ and ICC_3,2_, with corresponding 95% confidence limits, were calculated [26,27]. Shrout & Fleiss ICC model 3 (mixed model) was used, as this is recommended for establishing test-retest reliability [28]. ICC_3,1_ provides the reliability for a single measurement, whereas ICC_3,2_ provides reliability for the average of two measurements. Bland-Altman plots were created for each variable. The standard error of measurement (SEM) was calculated as the square root of the mean square error from repeated measures analysis of variance; this method calculates SEM independent of the ICC [29]. The minimum detectable change (MDC) was calculated as [29]:

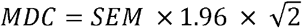

The number of trials needed to average together to achieve an ICC of at least 0.9 (*k*) was calculated using the Spearman-Brown formula [30]:

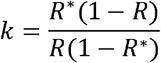

where *R* is the calculated ICC_3,1_ and *R*^*\**^ is the target ICC (0.9).

### Sample size justification

With 2 observations per participant, a sample size of >7 is sufficient to detect a minimum acceptable ICC of 0.4, and ideal reliability of 0.9, with α=0.05 and β=0.20 [31]. ICC of 0.4 suggests ‘fair’ reliability [32], while ICC of 0.9 is recommended for applying a measure to clinical decision making [28]. We included 20 participants to ensure wider variability in participant characteristics than would be achieved with a sample size of 7.

## RESULTS

Results of the analysis are presented in Table 2. ICC_3,1_ were between 0.62 (between-limb synchronization) and 0.91 (RMS of ML COP displacement). ICC_3,2_ were between 0.79 (displacement symmetry index) and 0.95 (RMS of ML COP displacement). The lower bound of the ICC_3,2_ confidence intervals were >0.50 for all variables. Bland-Altman plots (Figure 1) revealed possible proportional biases for RMS of ML COP displacement and velocity and between-limb synchronization, with larger differences between observations for participants with worse values (i.e., higher RMS of ML COP displacement or velocity, or lower between-limb synchronization).

**Table 2:** Means and reliabilities for each variable. Values presented for the three trials are means with standard deviations in parentheses. The intraclass correlations (ICCs) are presented with 95% confidence intervals in brackets.

|  | Trial 1 | Trial 2 | ICC<sub>3,1</sub> | ICC<sub>3,2</sub> | SEM | MDC | k |
| --- | --- | --- | --- | --- | --- | --- | --- |
| RMS of ML COP displacement (mm) | 6.5 (8.2) | 7.0 (9.4) | 0.91 [0.78, 0.96] | 0.95 [0.88, 0.98] | 2.7 | 7.5 | 1 |
| RMS of AP COP displacement (mm) | 6.5 (2.9) | 6.3 (2.8) | 0.82 [0.61, 0.93] | 0.90 [0.75, 0.96] | 1.2 | 3.3 | 2 |
| RMS of ML COP velocity (mm/s) | 17.7 (12.9) | 15.5 (8.8) | 0.80 [0.57, 0.92] | 0.89 [0.72, 0.96] | 4.9 | 13.6 | 3 |
| RMS of AP COP velocity (mm/s) | 21.0 (9.9) | 20.4 (7.0) | 0.82 [0.59, 0.92] | 0.90 [0.74, 0.96] | 3.7 | 10.1 | 3 |
| Displacement symmetry index | 0.41 (0.11) | 0.43 (0.12) | 0.66 [0.32, 0.85] | 0.79 [0.49, 0.92] | 0.07 | 0.18 | 5 |
| Velocity symmetry index | 0.41 (0.11) | 0.42 (0.12) | 0.68 [0.35, 0.86] | 0.81 [0.51, 0.92] | 0.04 | 0.10 | 5 |
| Between-limb synchronization | 0.76 (0.19) | 0.71 (0.24) | 0.62 [0.39, 0.87] | 0.83 [0.57, 0.93] | 0.13 | 0.35 | 6 |
| Sample entropy | 0.403 (0.071) | 0.409 (0.103) | 0.80 [0.57, 0.92] | 0.89 [0.72, 0.96] | 0.039 | 0.109 | 3 |

AP = antero-posterior; COP = centre of pressure; k = number of trials needed to average together to achieve $ICC_{3,k} \geq 0.9$ (rounded up); MDC = minimal detectable change; ML = medio-lateral; RMS = root mean square; SEM = standard error of measurement.

**Figure 1:**
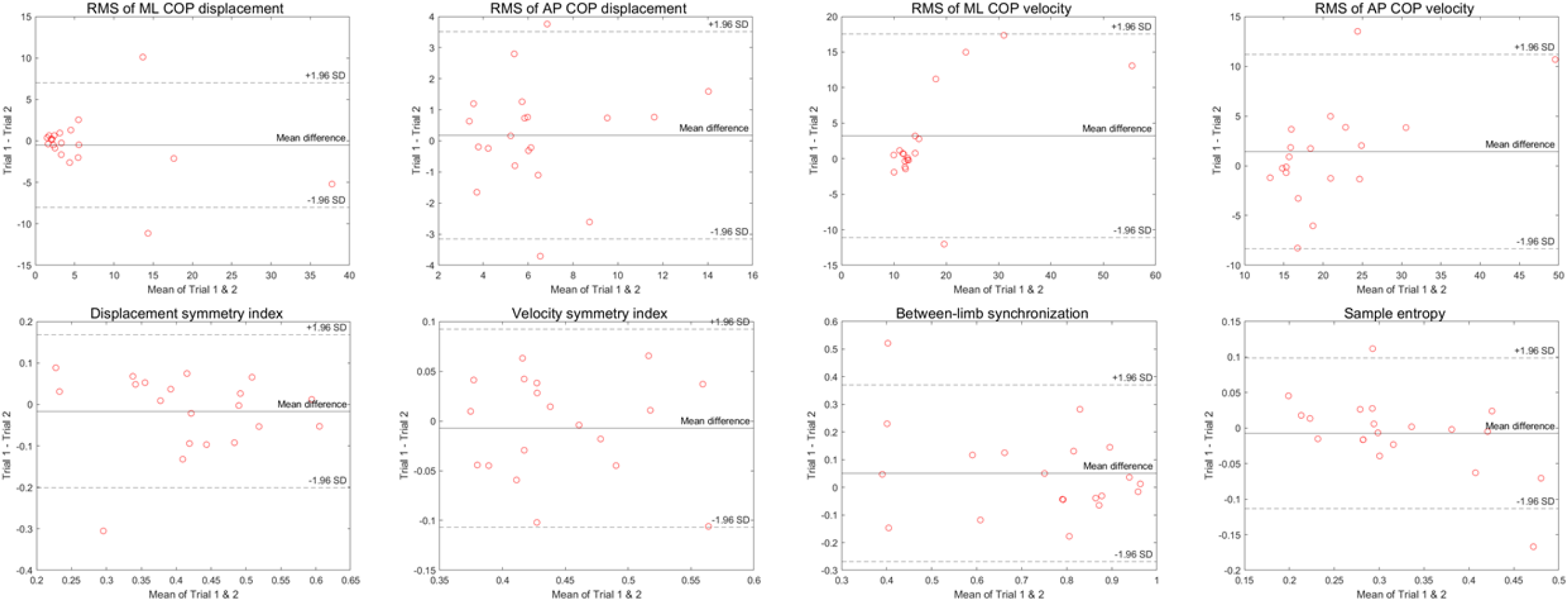
Bland-Altman plots. The x-axis on each plot is the mean of Trial 1 and Trial 2, and the y-axis is the difference between Trial 1 and Trial 2. Proportional biases are evident for RMS of ML COP displacement, RMS of ML COP velocity, and between-limb synchronization, with larger between-trial differences for participants with worse values (i.e., higher RMS of ML COP displacement and velocity, or lower between-limb synchronization). AP = antero-posterior; COP = centre of pressure; ML = medio-lateral; RMS = root mean square.

## DISCUSSION

This study aimed to determine the test-retest reliability of novel measures of quiet standing balance control in people with chronic stroke. We calculated reliabilities for a single 30-second trial (ICC_3,1_) and the mean of two trials (ICC_3,2_). Estimated ICC_3,2_ were all above the 0.75 threshold that most authors consider ‘good’ [28,33] or ‘excellent’ [32] reliability. Likewise, estimated ICC_3,1_ were above this threshold for all measures, except the symmetry indices and between-limb synchronization; ICC_3,1_ of 0.62-0.68 for symmetry indices and between-limb synchronization suggest ‘moderate’ [28,33] or ‘fair to good’ [32] reliability. Therefore, a single 30-second trial may be sufficient for using these measures in research studies [28]. However, higher reliabilities (> 0.9) are recommended for clinical decision making [28]. Only ICC_3,1_ for RMS of ML COP displacement, and ICC_3,2_ for RMS of ML and AP COP displacement and RMS of AP COP velocity met this higher threshold, with RMS of ML COP velocity and sample entropy almost meeting this threshold (ICCs = 0.89). Therefore, for clinical application of these measures, we recommend taking the average of 2 to 6 trials, rather than using a single trial.

In quiet standing, the COP moves to control postural sway [6]. Therefore, the magnitude of COP fluctuations can be used as a proxy measure for the amount of postural sway [34]. Previous studies have used measures based on variability and velocity of COP to quantify the magnitude of COP fluctuations, with velocity-based measures typically being more reliable than displacement-based measures [32]. In our study, we included RMS of both COP displacement and velocity to inform whether the symmetry index should be based upon RMS of COP displacement or velocity. Contrary to other studies, we found that RMS of COP displacement and velocity had similar reliabilities. However, the MDCs as a proportion of the sample mean were high for all measures, and the MDC exceeded the sample mean for RMS of COP displacement. Within our sample, RMS values were right-skewed; e.g., most participants had RMS of ML COP displacement around 5 mm or lower, whereas 4 participants had values of 10-40 mm (see Figure 1). Theoretically, RMS of COP has a minimum value of 0 and no maximum value. However, a value of 0 would be physiologically impossible in living humans; there will always be at least a small amount of postural sway that has to be controlled. Likewise, the range of COP movement is limited by the size of the base of support. Therefore, there are functional maxima and minima for RMS of COP measures, and it is likely that many participants in our study experienced a floor effect for these measures. For reference, 15 of our participants also reached an effective ceiling on the Berg Balance Scale, scoring the maximum value or within the MDC [35] of the maximum value. Therefore, the MDCs that we have calculated would only apply to participants who have RMS of COP that exceeds a ‘normal’ range, whereas participants within the ‘normal’ range have essentially no room for improvement.

To our knowledge, only one other study has attempted to determine the reliability of between-limb synchronization post-stroke [9]. Our ICCs were higher and MDC was lower than this previous study (within session ICC_3,1_: 0.18 and 0.57, MDC: 0.47 [9]). The previous study included people in both the sub-acute and chronic stages of stroke recovery. It is possible that balance measures are less stable, and therefore less reliable, among people earlier after stroke. For example, the Berg Balance Scale has a reportedly lower ICC (0.92 versus 0.95) and higher MDC (6.9 versus 4.7) for people with sub-acute compared to chronic stroke [35,36]. Both our study and this previous study reported a high MDC relative to the range of the scale; MDC were 0.35 (our study) and 0.47 (previous study) [9], whereas between-limb synchronization values can range from -1 to 1. However, it is likely that this measure is on a non-linear scale. That is, an increase in between-limb synchronization from 0 to 0.35 may be less of an improvement than an increase from 0.55 to 0.9. Similarly, it is possible that all values <0 are essentially equivalent, reflecting unsynchronized balance control; that is, an increase from -0.35 to 0 might not reflect any improvement in between-limb synchronization, meaning that this measure has a functional range of 0 to 1. If this is the case, an MDC of 0.35, or 35% of the functional range of the scale, means that very large improvements need to be made to exceed measurement error. Future work should provide further insights on clinical interpretation of specific values of this measure, and determine if MDC differs by baseline value.

To our knowledge, no previous study has aimed to determine the test-retest reliabilities of symmetry indices in people with chronic stroke. We calculated symmetry indices based on RMS of both COP displacement and velocity. Previous studies have calculated the symmetry index using RMS of COP displacement [37]; however, as velocity-based COP measures are usually more reliable than displacement-based measures [32], we also determined the reliability of the symmetry index using RMS of COP velocity. While ICCs were similar for both symmetry indices, the SEM, and consequently the MDC, was lower for the velocity symmetry index than the displacement symmetry index. Therefore, we recommend using the velocity symmetry index.

Others have aimed to determine the test-rest reliability of sample entropy of the COP among male youth athletes [38] and people with musculoskeletal disorders [39]. These studies reported within-session ICCs of 0.53-0.68 (approximately 1 minute break between trials [39]) and between-session ICCs of 0.76-0.81 [38,39], which were slightly lower than our study. Both of these studies calculated sample entropy based on the AP and ML COP separately, whereas we calculated sample entropy from the resultant COP signal [16]; this difference in signal processing may explain the higher ICCs observed in our study. Values of sample entropy can be influenced by choice of input parameters [16], which are typically determined for each specific dataset [24]. It is unknown how choice of input parameters might influence test-retest reliability of sample entropy measures.

Our study used relatively short trial durations (30 sec). Previous studies have found that increasing the trial duration can improve the reliability of COP-based balance measures [32,40]. However, in our clinical experience, fatigue and attentional difficulties can present challenges for including longer quiet standing balance trials for people with stroke [5]. Therefore, taking the average of several shorter trials may be preferable to including just one or two longer trials for improving accuracy of balance assessment post-stroke [32].

Our study is limited by inclusion of a relatively ‘high functioning’ sample of participants. Participants needed to be able to stand and walk without a gait aid to be eligible for the larger study. This resulted in many participants achieving a functional maximum or minimum on some measures. Therefore, the results of this study might not generalize to people with greater balance dyscontrol. Our study findings might also not generalize to other quiet standing balance testing conditions (e.g., eyes closed).

## CONCLUSIONS

The novel and stroke-specific measures of quiet standing balance analysed in this study, including symmetry indices, between-limb synchronization, and sample entropy, demonstrated ‘moderate’ to ‘excellent’ reliabilities. A single 30-second trial may have sufficient reliability for some research applications. However, for clinical applications, the average of at least 2 to 6 trials should be taken.

## Data Availability

The participants of this study did not give written consent for their data to be shared publicly, so supporting/raw data is not available.

## REFERENCES

1. Mansfield A, Inness EL, McIlroy WE. Stroke. In: Day BL, Lord SR, eds. Handbook of Clinical Neurology: Balance, Gait, and Falls. Vol 159. San Diego: Elsevier BV; 2018:205–228.

2. Jette DU, Halbert J, Iverson C, Micheli E, Shah P. Use of standardized outcome measures in physical therapist practice: perceptions and applications. Phys Ther. 2009;89:125–135.

3. Berg K, Wood-Dauphinée S, Williams JI, Gayton D. Measuring balance in the elderly: preliminary development of an instrument. Physiother Can. 1989;41:304–311.

4. Sibley KM, Straus SE, Inness EL, Salbach NM, Jaglal SB. Balance assessment practices and use of standardized measures among Ontario physical therapists. Phys Ther. 2011;91:1583–1591.

5. Mansfield A, Inness EL. Force plate assessment of quiet standing balance control: perspectives on clinical application within stroke rehabilitation. Rehabilitation Process and Outcome. 2015;4:7–15.

6. Winter DA. Human balance and posture control during standing and walking. Gait Posture. 1995;3:193–214.

7. Gasq D, Labrunée M, Amarantini D, Dupui P, Montoya R, Marque P. Between-day reliability of centre of pressure measures for balance assessment in hemiplegic stroke patients. J Neuroeng Rehabil. 2014;11:39.

8. Gray VL, Ivanova TD, Garland SJ. Reliability of center of pressure measures with and between sessions in individuals post-stroke and healthy controls. Gait Posture. 2014;40:198–203.

9. Martello SK, Boumer TC, deAlmeida JC, et al. Reliability and minimal detectable change of between-limb synchronization, weight-bearing symmetry, and amplitude of postural sway in individuals with stroke. Research on Biomedical Engineering. 2017;33:113–120.

10. Winter DA, Prince F, Stergiou P, Powell C. Medial-lateral and anterior-posterior motor responses associated with centre of pressure changes in quiet stance. Neurosci Res Commun. 1993;12:141–148.

11. Mansfield A, Danells CJ, Inness EL, Mochizuki G, McIlroy WE. Between-limb synchronization for control of standing balance in individuals with stroke. Clin Biomech. 2011;26:312–317.

12. Roerdink M, Geurts ACH, de Haart M, Beek PJ. On the relative contribution of the paretic leg to the control of posture after stroke. Neurorehabil Neural Repair. 2009;23:267–274.

13. Genthon N, Rougier P, Gissot A-S, Froger J, Pélissier J, Pérennou DA. Contribution of each lower limb to upright standing in stroke patients. Stroke. 2008;39:1793–1799.

14. Mansfield A, Wong JS, McIlroy WE, et al. Do measures of reactive balance control predict falls in people with stroke returning to the community? Physiotherapy. 2015;101:373–380.

15. Roerdink M, De Haart M, Daffertshofer A, Donker SF, Geurts AC, Beek PJ. Dynamical structure of center-of-pressure trajectories in patients recovering from stroke. Exp Brain Res. 2006;174:256–269.

16. Roerdink M, Hlavackova P, Vuillerme N. Center-of-pressure regularity as a marker for attentional investment in postural control: a comparison between sitting and standing postures. Hum Mov Sci. 2011;30:190–202.

17. Mansfield A, Aqui A, Danells CJ, et al. Does perturbation-based balance training prevent falls among individuals with chronic stroke? A randomised controlled trial. BMJ Open. 2018;8:e021510.

18. Schinkel-Ivy A, Huntley AH, Aqui A, Mansfield A. Does perturbation-based balance training improve control of reactive stepping in individuals with chronic stroke. J Stroke Cerebrovasc Dis. 2019;28:935–943.

19. Goldstein LB, Bertels C, Davis JN. Interrater reliability of the NIH Stroke Scale. Arch Neurol. 1989;46:660–662.

20. Gowland C, Stratford P, Ward M, et al. Measuring physical impairment and disability with the Chedoke-McMaster Stroke Assessment. Stroke. 1993;24:58–63.

21. Podsiadlo D, Richardson S. The Timed “Up & Go”: A test of basic functional mobility for frail elderly persons. J Am Geriatr Soc. 1991;39:142–148.

22. McIlroy WE, Maki BE. Preferred placement of the feet during quiet stance: development of a standardized foot placement for balance testing. Clin Biomech. 1997;12:66–70.

23. Goldberger AL, Amaral LAN, Glass L, et al. PhysioBank, PhysioToolkit, and PhysioNet: components of a new research resource for complex physiologic signals. Circulation. 2000;101:e215–220.

24. Lake DE, Richman JS, Griffin MP, Moorman JR. Sample entropy analysis of neonatal heart rate variability. Am J Physiol Regul Integr Comp Physiol. 2002;283:R789–797.

25. Hammer RM. Compute six intraclass correlation measures. 2000; http://support.sas.com/kb/25/031.html. Accessed 3 Dec 2012.

26. Shrout PE, Fleiss JL. Intraclass correlations: uses in assessing rater reliability. Psychol Bull. 1979;86:420–428.

27. Hallgreen KA. Computing inter-rater reliability for observational data: an overview and tutorial. Tutor Quant Methods Psychol. 2012;8:23–34.

28. Portney L, Watkins M. Foundations of Clinical Research. Vol 3rd. Philadelphia: F. A. Davis Company; 2015.

29. Weir JM, Vincent W. Statistics in Kinesiology. 5th ed. Champaign, IL: Human Kinetics; 2021.

30. Lafond D, Corriveau H, Hebert R, Prince F. Intrasession reliability of center of pressure measures of postural steadiness in healthy elderly people. Arch Phys Med Rehabil. 2004;85:896–901.

31. Walter SD, Eliasziw M, Donner A. Sample size and optimal designs for reliability studies. Stat Med. 1998;17:101–110.

32. Ruhe A, Fejer R, Walker B. The test-retest reliability of centre of pressure measures in bipedal stance task conditions - a systematic review of the literature. Gait Posture. 2010;32:436–445.

33. Koo TK, Li MY. A guideline of selecting and reporting intraclass correlation coefficients for reliability research. J Chiropr Med. 2016;15:155–163.

34. de Haart M, Geurts AC, Huidekoper SC, Fasotti L, van Limbeek J. Recovery of standing balance in postacute stroke patients: a rehabilitation cohort study. Arch Phys Med Rehabil. 2004;85:886–895.

35. Hiengkaew V, Jitaree K, Chaiyawat P. Minimal detectable changes of the Berg Balance Scale, Fugl-Meyer Assessment Scale, Timed “Up & Go” Test, gait speeds, and 2-minute walk test in individuals with chronic stroke with different degrees of ankle plantarflexor tone. Arch Phys Med Rehabil. 2012;93:1201–1208.

36. Stevenson TJ. Detecting change in patients with stroke using the Berg Balance Scale. Aust J Physiother. 2001;47:29–38.

37. Rougier PR, Genthon N. Dynamical assessment of weight-bearing asymmetry during upright quiet stance in humans. Gait Posture. 2009;29:437–443.

38. Campolettano ET, Madigan ML, Rowson S. Reliability of center of pressure-based measures during dual-task postural control testing in a youth population. Int J Sports Phys Ther. 2020;15:1036–1043.

39. Mazaheri M, Negahban H, Salavati M, Sanjari MA, Parnianpour M. Reliability of recurrence quantification analysis measures of the center of pressure during standing in individuals with musculoskeletal disorders. Med Eng Phys. 2010;32:808–812.

40. Carpenter MG, Frank JS, Winter DA, Peysar GW. Sampling duration effects on centre of pressure summary measures. Gait Posture. 2001;13:35–40.

